# Telemedicine to Address Inequities in Access to Primary Health Care for Minority Groups: A protocol for a Systematic Review and Meta-Analysis

**DOI:** 10.1101/2024.11.27.24317907

**Authors:** Gabriela de Oliveira Laguna Silva, Gabriel Ricardo Fernandes, Emmanuelle Moreira Pereira, Jacqueline Castro da Rocha, Gabriela Tizianel Aguilar, Tiago Sigal Linhares, Andressa Dutra Dode, Felipe Cezar Cabral, Taís de Campos Moreira, Maria Eulália Vinadé Chagas

## Abstract

**Background:** The present article outlines the protocol for a systematic review and meta-analysis aimed at consolidating the available evidence on how telemedicine can reduce inequities in access to Primary Health Care (PHC) among minority populations. The review seeks to identify barriers, facilitators, and factors that influence the effectiveness of this intervention across different settings.

**Methods:** This protocol for a systematic review and meta-analysis is reported in line with Prisma-P and the results will be reported using PRISMA-E flowchart. The eligibility criteria was defined using the PICOS approach. Population is minority groups that have used telemedicine, the intervention is synchronous and asynchronous telemedicine, the comparator will consist of minority groups that have received usual care, outcome is access to primary health care and we will include randomized controlled trials. We will search MEDLINE (via PubMed), Scopus, Latina American and Caribbean Health Sciences Literature (LILACS) databases. The risk of bias will be assessed using the ROBINS-I and the confidence in cumulative evidence will be assessed using the GRADE equity tool.

**Discussion:** It is expected to provide pragmatic results, such supporting decision-making regarding the creation and implementation of new public policies, the development of clinical guidelines, and the optimization of resources aimed at equitable access.

**Systematic review registration:** PROSPERO CRD42024581305

## Background

Primary Health Care (PHC) is fundamental to the health system, acting as the initial point of contact for individuals with health services and playing a vital role in advancing health equity^1^. As one of the pillars of healthcare systems, PHC provides continuous, comprehensive, and accessible care, which is essential not only for disease prevention and health promotion but also for the management of chronic conditions^1^. However, despite its importance, minority groups in various regions face significant barriers that limit their access to these essential services. These health inequities are often reflected in disparities in social determinants of health (SDH) such as place of residence, race/ethnicity, occupation, gender, religion, education, socioeconomic status, and social capital framework ^2,3^.

In low and middle-income countries (LMIC), these inequities are particularly pronounced, where many populations face severe barriers to accessing quality healthcare services^2,4^. For instance, in rural or remote areas, for instance, the shortage of healthcare professionals and the distance to health care centers make access to Primary Health Care (PHC) even more challenging. Additionally, economic and social factors exacerbate these disparities, leading to worse health outcomes for these populations^3,4^.

In this context, telemedicine has emerged as a promissing tool to mitigate healthcare access, offering viable solutions to overcome geographical and socioeconomic barriers by providing remote healthcare services via digital technologies^1^. This is especially relevant in rural areas, where physical access to healthcare is limited. Furthermore, telemedicine has the potential to reduce linguistic and cultural barriers, improving communication and adherence to treatment among patients^2^.

As telemedicine becomes increasingly relevant on the global stage, it is crucial to understand its impact on health inequities and how it can be optimized to benefit the most vulnerable populations^1,5^. Marginalized populations, such as Indigenous populations, LGBTQIAP+ individuals, homeless populations, people with disabilities, Quilombolas populations, Black populations, incarcerated populations, and those reentering society from the prison system, face even greater barriers to accessing healthcare. These populations often do not benefit equally from technological advancements in health due to the stratifying social forces present in societies^2,4,6^.

The Pan American Health Organization (PAHO) highlights that telemedicine can improve access and timeliness in the delivery of healthcare services, particularly benefiting populations that face significant challenges in accessing these services^5^. Telemedicine should be seen as a pathway. One of the greatest challenges in its evaluation is the lack of knowledge and proper use, which limits the development of reliable indicators. Given the current shortage of evaluation tools, it is essential that assessments focus on demonstrating the comparative advantages of telemedicine over traditional healthcare practices that do not incorporate technology^7^. While telemedicine is widely promoted as a means to enhance healthcare access, evidence of its effectiveness in reducing health inequities remains fragmented and sparse^1,6^. As a result, the current synthesis of evidence reveals significant gaps in understanding how to effectively address these inequities.^3^.

This review is necessary to explore the impact of telemedicine across various contexts, especially for populations that have historically encountered substantial barriers to healthcare access. The goal is to consolidate the available evidence on how telemedicine can help reduce inequities in access to Primary Health Care (PHC) among minority populations, identifying barriers, facilitators, and factors that affect the effectiveness of this intervention in different settings.

## Methods

This protocol for a systematic review and meta-analysis is reported in line with Preferred Reporting Items for Systematic Reviews and Meta-analysis Protocols (PRISMA-P)^8^ (Supplemental material 1). Details of this protocol were registered in the International Prospective Register of Systematic Reviews (PROSPERO) and can be accessed at PROSPERO^9^. We aimed to conclude and publish this systematic review in the first semester of 2025. Any changes to the protocol will be made on the PROSPERO.

### Eligibility criteria

The eligibility criteria was defined using Population, Intervention, Comparator, Outcomes and Study design (PICOS) approach.

### Population

We will consider studies on minority group, as:

1. indigenous population
2. LGBTQIAP+ population (Lesbian, Gay, Bi, Trans, Queer, intersex, asexual/agender/aromantic, Pan/Poly, Non-binary, and more)
3. Homeless population
4. People with disabilities
5. Quilombola population
6. Black population
7. Incarcerated population
8. Formerly incarcerated individuals

### Intervention

The intervention will be the use of synchronous and asynchronous telemedicine.

### Comparators

The comparator will consist of minority groups that have received usual care (face-to-face consultation).

### Outcomes

We will consider studies that bring information about access to primary health care.

### Studies design

We will include randomized controlled trials.

### Report characteristics

There will be no language or date restrictions. Gray literature and conference abstracts will be excluded.

### Information sources

We will search the following electronic bibliographic databases: MEDLINE (via PubMed), Scopus, Latina American and Caribbean Health Sciences Literature

(LILACS). The references of the selected articles will be manually screened to identify potential studies that were not retrieved by the search strategy.

### Search strategy

The search strategy was developed in collaboration with a librarian to ensure it was comprehensive and accurate. First, the strategy was designed for the PubMed database and, after being reviewed and approved, it was adapted for other databases, Scopus and LILACS. The strategy was structured according to the PICO framework, and to capture populations exposed to social determinants of health but not explicitly identified as minority groups, we incorporated the terms “minority group” AND “inequities.” Boolean operators, indexed database terms, and free-text terms were utilized to optimize the search for relevant studies across all targeted databases (Supplemental Material 2). This comprehensive approach ensured that all pertinent literature was included, broadening the scope of the review to encompass studies addressing both minority groups and inequities related to healthcare access.

### Study records

#### Data management

The records retrieved from the databases will be imported into Rayyan©, an online systematic review tool^10^. Reviewers will have access to a shared Rayyan© account so that they can read the abstracts of retrieved studies.

#### Selection process

Two groups of independent reviewers will be randomly formed (Group 1: EP, GF, AD e MC e Group 2: GL, GT, TL e JR) to assess the titles, keywords and abstracts of the retrieved articles using Rayyan. If the eligibility cannot be determined, the articles will be classified as eligible for the full-text review stage. A third reviewer (TM) will be responsible for reaching consensus on articles with discrepant evaluations between the reviewer groups.

#### Data collection process

The relevant information will be collected independently by both groups of reviewers from each eligible study and transcribed into a Google Sheets spreadsheet.

### Data items

The information that will be extracted from the eligible studies and transcribed into a Google Sheets spreadsheet includes: year of publication, country where the study was conducted, study characteristics, sample size, demographic information of participants, information of PROGRESS-Plus to identify the social determinants of health, telemedicine characteristics, results, and conclusions.

### Outcomes and prioritization

The primary outcome is the access to primary health care, defined as the individual’s ability to obtain basic and essential health services that meet their health needs, which includes availability, accessibility, accommodation, affordability and acceptability^11^. The additional outcomes are the quality/effectiveness of access for users from low- and middle-income countries compared to developed countries, and the impact of telemedicine overuse on clinical outcomes.

### Risk of bias in individual studies

Two groups of independent reviewers (Group 1: EP, GF, AD e MC e Group 2: GL, GT, TL e JR) will assess the risk of bias using the ROBINS-I^12^ tool in non-randomized Studies of interventions.

### Data synthesis

The study selection description will be done using a PRISMA-E^13^ flowchart. All included studies will have qualitative and quantitative data related to the research question extracted and synthesized. If there is sufficient and appropriate data, metaanalyses will be conducted for the groups of interest.

### Meta-bias(es)

Funnel plots will be generated and visually examined for asymmetry, which may suggest the presence of publication bias^14^. When the meta-analysis includes 10 or more studies, the Egger test will be performed to assess funnel plots asymmetry^14^.

### Confidence in cumulative evidence

Two groups of independent reviewers (Group 1: EP, GF, AD e MC e Group 2: GL, GT, TL e JR) will independently assess the quality of evidence for all outcomes using the Grading of Recommendations Assessment, Development and Evaluation (GRADE) equity approach^15^.

Conclusions regarding GRADE for each outcome will be presented in a Summary of Findings table. An overall GRADE will be assigned to the set of all outcomes.

## Discussion

This article is a protocol to guide a systematic review which will synthesize the existing literature on access to the primary health care for minority groups through telemedicine. It aims to provide pragmatic outcomes, such as supporting decision-making regarding the creation and implementation of new public policies, the development of clinical guidelines, and resource optimization to promote equitable access. It is understood that some limitations may arise throughout the work, such as insufficient studies for certain minority groups. However, there are several relevant aspects, such as the use of an established methodology and guidelines in conducting the study, the publication of the current protocol, and registration in PROSPERO, aiming for transparency and reproducibility. The results will be published in a peer-reviewed journal.

## Data Availability

Not applicable.

## Acknowledgments

Francieli Ariane Lehnen Muck, librarian at FAMED/UFRGS, for her support in the literature search.

