## Supplemental material 2 for "Telemedicine to Address Inequities in Access to Primary Health Care for Minority Groups: A protocol for a Systematic Review and Meta-Analysis"

**Acknowledgments:** Francieli Ariane Lehnen Muck, librarian at FAMED/UFRGS, for her support in the literature search.

**Conflict of Interest:**The authors declare that there is no conflict of interest.

**Funding:** Provided by the Brazilian Ministry of Health, through the Institutional Development Program of the Brazilian National Health System (PROADI-SUS).

**Systematic review registration:** PROSPERO CRD42024581305

**Supplemental material 2**

PubMed:

(Telemedicine[mh] OR Telemedicine[tiab] OR Virtual Medicine[tiab] OR Tele Referral*[tiab] OR Mobile Health[tiab] OR mHealth[tiab] OR Telehealth[tiab] OR eHealth[tiab] OR Telecare[tiab] OR Tele Care[tiab])

AND

(Primary Health Care[mh] OR Primary Health Care[tiab] OR Primary Care[tiab] OR Primary Healthcare[tiab])

AND

(Indigenous Peoples[mh] OR Indigenous[tiab] OR First Nation People*[tiab] OR Tribes[tiab] OR Native*[tiab] OR Sexual and Gender Minorities[mh] OR Non Heterosexual*[tiab] OR Sexual Dissident*[tiab] OR GLBT[tiab] OR LGBT*[tiab] OR Gender Diverse[tiab] OR LBG Person*[tiab] OR Lesbigay[tiab] OR Sexual Minorit*[tiab] OR Bisexual*[tiab] OR Homosexual*[tiab] OR Queer*[tiab] OR Gays[tiab] OR Gay[tiab] OR Lesbian*[tiab] OR Gender Minorit*[tiab] OR Ill-Housed Persons[mh] OR Ill Housed Person*[tiab] OR Shelterless Person*[tiab] OR Homeless Person*[tiab] OR Unhoused Person*[tiab] OR Street People[tiab] OR Homelessness[tiab] OR Homeless Shelter*[tiab] OR Disabled Persons[mh] OR Disabled Person*[tiab] OR People with Disabilit*[tiab] OR Handicapped[tiab] OR Physically Disabled[tiab] OR Physically Challenged[tiab] OR Quilombola[tiab] OR Black People[mh] OR Black*[tiab] OR Negro*[tiab] OR African Continental Ancestry Group[tiab] OR African American*[tiab] OR Afro American*[tiab] OR Prisoners[mh] OR Prisoner*[tiab] OR Incarcerated Individual*[tiab] OR Imprisoned Individual*[tiab] OR Inmate*[tiab] OR Hostage*[tiab] OR Detained Person*[tiab] OR Ex-Prisoner*[tiab])

AND

(Health Inequities[mh] OR Health Equity[mh] OR Inequit*[tiab] OR Inequalit*[tiab] OR Disparit*[tiab] OR Equit*[tiab] OR Equalit*[tiab] OR Disadvantage*[tiab] OR Residence Characteristics[mh] OR Residence[tiab] OR Place of Birth[tiab] OR Birth Place[tiab] OR Communit*[tiab] OR Residential[tiab] OR Domicile*[tiab] OR Racial Groups[mh] OR Racial Group*[tiab] OR Race*[tiab] OR Ethnicity[mh] OR Ethnicity[tiab] OR Ethnic[tiab] OR Culture[mh] OR Culture[tiab] OR Customs[tiab] OR Language[mh] OR Language[tiab] OR Dialect*[tiab] OR Employment[mh] OR Employment[tiab] OR Occupational Status[tiab] OR Labor Force*[tiab] OR Underemployment[tiab] OR occupation*[tiab] OR Gender Identity[mh] OR Gender[tiab] OR Religion[mh] OR Religio*[tiab] OR Prayer*[tiab] OR Education[mh] OR Educat*[tiab] OR Socioeconomic Factors[mh] OR Social and Economic Factors[tiab] OR Economic and Social Factors[tiab] OR Socioeconomic[tiab] OR socio-economic[tiab] OR High Income Population*[tiab] OR Land Tenure[tiab] OR Standard of Living[tiab] OR Living Standard*[tiab] OR Social Capital[mh] OR Social Capital[tiab] OR Social Infrastructure*[tiab])

Scopus:

TITLE-ABS-KEY(Telemedicine OR "Virtual Medicine" OR "Tele Referral*" OR "Mobile Health" OR mHealth OR Telehealth OR eHealth OR Telecare OR "Tele Care")

AND

TITLE-ABS-KEY("Primary Health Care" OR "Primary Care" OR "Primary Healthcare")

AND

TITLE-ABS-KEY(Indigenous OR "First Nation People*" OR Tribes OR Native* OR "Non Heterosexual*" OR "Sexual Dissident*" OR GLBT OR LGBT* OR "Gender Diverse" OR "LBG Person*" OR Lesbigay OR "Sexual Minorit*" OR Bisexual* OR Homosexual* OR Queer* OR Gays OR Gay OR Lesbian* OR "Gender Minorit*" OR "Ill Housed Person*" OR "Shelterless Person*" OR "Homeless Person*" OR "Unhoused Person*" OR "Street People" OR Homelessness OR "Homeless Shelter*" OR "Disabled Person*" OR "People with Disabilit*" OR Handicapped OR "Physically Disabled" OR "Physically Challenged" OR Quilombola OR Black* OR Negro* OR "African Continental Ancestry Group" OR "African American*" OR "Afro American*" OR Prisoner* OR "Incarcerated Individual*" OR "Imprisoned Individual*" OR Inmate* OR Hostage* OR "Detained Person*" OR "Ex-Prisoner*")

AND

TITLE-ABS-KEY(Inequit* OR Inequalit* OR Disparit* OR Equit* OR Equalit* OR Disadvantage* OR Residence OR "Place of Birth" OR "Birth Place" OR Communit* OR Residential OR Domicile* OR "Racial Group*" OR Race* OR Ethnicity OR Ethnic OR Culture OR Customs OR Language OR Dialect* OR Employment OR "Occupational Status" OR "Labor Force*" OR Underemployment OR occupation* OR Gender OR Religio* OR Prayer* OR Educat* OR "Social and Economic Factors" OR "Economic and Social Factors" OR Socioeconomic OR "socio-economic" OR "High Income Population*" OR "Land Tenure" OR "Standard of Living" OR "Living Standard*" OR "Social Capital" OR "Social Infrastructure*")

Lilacs:

(mh:(H02.403.840* OR L01.462.500.847.652* OR N04.590.374.800* OR SP2.840.065.715* OR SP2.840.566*) OR tw:(Telemedicin* OR "Virtual Medicine" OR "Tele Referral" OR "Tele Referrals" OR "Mobile Health" OR mHealth OR Telehealth OR eHealth OR Telecare OR "Tele Care" OR "Ciber Saúde" OR Cibersaúde OR "Medicina 2.0" OR "Medicina Virtual" OR "Saúde 2.0" OR "Saúde Eletrônica" OR "e-Saúde" OR eSaúde OR mSaúde OR uSaúde OR "Tele-Serviços em Saúde" OR Teleassistência OR Telecuidado OR Telecura OR Telereferenciação OR Telessaúde OR Telesserviços OR eSalud OR mSalud OR uSalud OR "Ciber Salud" OR Cibersalud OR "Salud 2.0" OR "Salud Electrónica" OR "Salud Móvil" OR Telesalud OR "Tele Referencia" OR Teleasistencia OR Teleservicio*))

AND

(mh:(N04.590.233.727* OR SP2.630.121*) OR tw:("Primary Health Care" OR "Primary Care" OR "Primary Healthcare" OR "Atendimento Básico" OR "Atendimento Primário" OR "Atenção Básica" OR "Atenção Primária" OR "Cuidado Primário" OR "Cuidado de Saúde Primário" OR "Cuidados Primários" OR "Cuidados de Saúde Primários" OR "Primeiro Nível de Assistência" OR "Primeiro Nível de Atendimento" OR "Primeiro Nível de Atenção" OR "Primeiro Nível de Cuidado" OR "Primeiro Nível de Cuidados" OR "Asistencia Primaria" OR "Atención Básica" OR "Atención Primaria" OR "Atención Sanitaria de Primer Nivel" OR "Primer Nivel de Asistencia" OR "Primer Nivel de Atención" OR "Primer Nivel de la Asistencia" OR "Asistencia Sanitaria de Primer Nivel"))

AND

(mh:(M01.270.968* OR SP3.522.561.162.267.602* OR M01.270.988* OR SP3.522.561.162.300.451* OR M01.325* OR M01.150* OR SP3.522.561.162.267.713* OR M01.686.372* OR M01.729*) OR tw:(Indigenous OR "First Nation People" OR "First Nation Peoples" OR Tribes OR Nativ* OR Indígena* OR Aborigen* OR "Non Heterosexual" OR "Non Heterosexuals" OR "Sexual Dissident" OR "Sexual Dissidents" OR GLBT OR LGBT* OR "Gender Diverse" OR "LBG Persons" OR "LBG Person" OR Lesbigay OR "Sexual Minority" OR "Sexual Minorities" OR Bisexual* OR Homosexual* OR Queer* OR Gays OR Gay OR Lesbian* OR "Gender Minorities" OR "Gender Minority" OR Homossex* OR Lésbic* OR Bissex* OR Transex* OR Intersexuais OR Assexuais OR "Minorias Sexuais" OR "Minorias de Gênero" OR "Pessoas LGB" OR "não Heterossexuais" OR Gais OR "Género Diverso" OR "Minorías Sexuales" OR "Personas LBG" OR "no Heterosexuales" OR "Ill Housed Person" OR "Ill Housed Persons" OR "Shelterless Person" OR "Shelterless Persons" OR "Homeless Person" OR "Homeless Persons" OR "Unhoused Person" OR "Unhoused Persons" OR "Street People" OR Homelessness OR "Homeless Shelter" OR "Homeless Shelters" OR "Morador de Rua" OR "Moradores de Rua" OR "Pessoas em Situação de Rua" OR "Pessoas sem Lar" OR "População em Situação de Rua" OR "Sem-Teto" OR "Personas en Situación de Calle" OR "Personas sin Hogar" OR "Personas sin Refugio" OR "Sin Techo" OR Sintecho OR "Disabled Person" OR "Disabled Persons" OR "People with Disability" OR "Persons with Disabilities" OR Handicapped OR "Physically Disabled" OR "Physically Challenged" OR Deficienc* OR Cadeirant* OR "Usuário de Cadeira de Rodas" OR Discapacidad* OR Quilomb* OR Cumbes OR Marroons OR Mocambos OR Palenques OR Black* OR Negr* OR "African Continental Ancestry Group" OR "African American" OR "African Americans" OR "Afro American" OR "Afro Americans" OR Afrodescendent* OR "Grupo com Ancestrais Africanos Continentais" OR "Grupo com Ancestrais do Continente Africano" OR "Grupo de Ancestralidade no Continente Africano" OR "Grupo de Ascendência Continental Africana" OR "Grupos Étnicos da África" OR "Ascendência Africana" OR Afrodescendient* "Etnias de África" OR "Grupo de Ancestro Africano Continental" OR "Grupo de Ascendencia Continental Africana" OR "Afro-Americano" OR "Afro-Americanos" OR "Africano Americano" OR "Africanos-Americanos" OR Afroamericano* OR Prisoner* OR "Incarcerated Individual" OR "Incarcerated Individuals" OR "Imprisoned Individual" OR "Imprisoned Individuals" OR Inmate* OR Hostage* OR "Detained Person" OR "Detained Persons" OR "Ex-Prisoner" OR Prisioneir* OR Cativ* OR Detent* OR Encarcerad* OR Preso* OR "Pessoa Privada de Liberdade" OR "Pessoas Privadas de Liberdade" OR "População Privada de Liberdade" OR Detid* OR Prisioneir* OR Refém OR Reféns OR Detenid* OR Encarcelad* OR "Persona Privada de Libertad" OR "Personas Privadas de Libertad" OR Reclus* OR Rehenes))

AND

(mh:(I01.240.425.513* OR N01.224.425.394* OR N04.590.374.350.500* OR N05.300.430.383* OR SP1.852.365* OR SP2.070.578* OR SP9.242.315.475.156.606* OR N01.224.791* OR N06.850.505.400.800* OR SP2.070.578.468.322* OR SP3.311.150* OR SP5.312.109.495.359* OR M01.686.477.625.594* OR M01.686.477.625.188* OR N01.224.317* OR SP3.522.561.162.267* OR I01.076.201.450* OR I01.880.853.100* OR SP3.311.900.898.314* OR F01.145.209.399* OR L01.559* OR SP3.311.900.353.100.548.300* OR N01.824.245* OR SP2.070.315.420.435.671* OR SP3.522.233.320* OR F01.393.446.250* OR F01.752.747.385.200* OR F01.752.747.722.200* OR F02.739.794.793.200* OR SP3.311.900.353.202.466* OR K01.844* OR SP2.070.315.420.754* OR SP3.311.900.898.314.587* OR I02* OR I01.880.853.996* OR N01.824* OR SP2.070.315.420.566* OR SP3.311.900.686.409* OR SP3.522.233* OR 01.880.853.375* OR SP3.311.750.298.366.120*) OR tw:(Inequit* OR Inequalit* OR Disparit* OR Equit* OR Equalit* OR Disadvantage* OR Desigualdad* OR Inequalidad* OR Iniquidad* OR Inequidad* OR Disparidad* OR Equidad* OR Residenc* OR "Place of Birth" OR "Birth Place" OR Communit* OR Residential OR Domicil* OR Habitação OR Comunidad* OR "Local de Nascimento" OR Localidade OR Naturalidade OR Vizinhanç* OR Vivienda OR "Lugar de Nacimiento" OR Vecindario* OR "Racial Group" OR "Racial Groups" OR Race* OR "Grupos Raciais" OR Raça* OR "Grupos Raciales" OR Raza* OR Ethnicity OR Ethnic OR Etnicid* OR Etnia* OR Étnic* OR Cultur* OR Customs OR Costume* OR Costumbre* OR Language OR Dialect* OR Idioma OR Dialeto OR Lenguaje OR Employment OR "Occupational Status" OR "Labor Force" OR "Labor Forces" OR Underemployment OR occupation* OR Emprego OR Ocupação OR "Status Laboral" OR "Status Ocupacional" OR Empleo OR "Ocupación Laboral" OR Gender OR Gênero OR Religi* OR Prayer* OR Educat* OR Educaç* OR "Social and Economic Factors" OR "Economic and Social Factors" OR Socioeconomic OR "socio-economic" OR "High Income Population" OR "High-Income Populations" OR "Land Tenure" OR "Standard of Living" OR "Living Standard" OR "Living Standards" OR "Fatores Econômicos e Sociais" OR "Fatores Sociais e Econômicos" OR "Factores Económicos y Sociales" OR "Factores Sociales y Económicos" OR "Social Capital" OR "Social Infrastructure" OR "Social Infrastructures" OR "Capital Social" OR "Infraestrutura Social" OR "Infrastructura Social"))

AND (db:("LILACS"))
